# Causal Effect of *Helicobacter Pylori* Infection on Coronary Heart Disease is mediated by Body Mass Index: A Mendelian Randomization Study

**DOI:** 10.1101/2023.03.08.23287014

**Authors:** Bing Li, Yaoting Zhang, Yang Zheng, He Cai

**Affiliations:** Department of Cardiovascular Diseases, The First Hospital of Jilin University, Changchun China

**Author notes:** **Correspondence** Yang Zheng and He Cai are co-corresponding authors. Postal address: Department of Cardiovascular Diseases, The First Hospital of Jilin University No. 1 Xinmin Ave, Changchun, China 130021.

**Keywords:** mendelian randomization, helicobacter pylori, coronary heart disease, inflammation, body mass index

## Abstract

**Background:** More than half of the world’s population have been infected with *H. pylori*, however the relationship between *H. pylori* infection and coronary heart disease (CHD) is unknown.

**Methods:** This study used mendelian randomization (MR) analyses. The instrument variables for *H. pylori* infection were genetic variables (rs10004195 and rs368433) obtained from a published study. The outcome data included diagnosis, prognosis, and pathogenesis data for CHD, which were extracted from the public genome-wide association studies database, mainly from the CARDIoGRAMplusC4D consortium, UK Biobank, IEU database, and FinnGen database. MR analyses were performed per outcome database and were conducted by reverse analysis. Two step MR analyses were used to explore indirect pathogenic factors of *H. pylori* infection.

**Results:** Genetically-predicted *H. pylori* infection was causally associated with body mass index (BMI) (β, 0.022; 95% CI, 0.008–0.036; *p*-value = 0.001), but not with the diagnosis of CHD (OR, 0.991; 95%CI, 0.904–1.078; *p*-value = 0.842, IEU database; OR, 1.049; 95% CI, 0.980–1.118; *p*-value = 0.178, FinnGen database) and prognosis of CHD (OR, 0.999; 95% CI, 0.997–1.001; *p*-value = 0.391, IEU database; OR, 1.022; 95% CI, 0.922–1.123; *p*-value = 0.663, FinnGen database). The causal effect of *H. pylori* infection on CHD is mediated by BMI. Inverse MR showed no causal effect of CHD on *H. pylori* infection.

**Conclusions:** Our findings confirm the causal effect of *H. pylori* infection on CHD is mediated by BMI. Eradication or prevention of *H. pylori* infection may have a clinical benefit for patients with CHD indirectly.

**Clinical Perspective:** *What Is New?:*

- Genetically-predicted *H. pylori* infection was causally associated with body mass index, but not with the diagnosis and prognosis (major adverse cardiovascular events) of coronary heart disease.
- The causal effect of *H. pylori* infection on coronary heart disease is mediated by body mass index. Inverse mendelian randomization analyses showed no causal effect of coronary heart disease on *H. pylori* infection.

*What Are the Clinical Implications?:*

- Our findings confirm that the causal effect of H. pylori infection on coronary heart disease is partially mediated by body mass index.
- Eradication or prevention of H. pylori infection may have a clinical benefit for patients with CHD indirectly.

## Introduction

Coronary heart disease (CHD) is caused by atherosclerosis, including angina pectoris and myocardial infarction (MI), the leading cause of mortality in many countries. ^1^ The etiology, pathogenesis and prognosis of CHD are complex and have not been fully understood until now. *Helicobacter pylori* (*H. pylori)* is a Gram-negative bacterium that primarily inhabits the stomach and duodenum. ^2^ More than half of the world’s population have been infected with *H. pylori*.^3^ In addition to causing gastrointestinal diseases, ^4^ *H. pylori* can also induce some systemic reactions, such as abnormal glucose ^5^ and lipid metabolism, ^6^ blood hypercoagulability,^7, 8^ and chronic inflammatory reactions,^9-11^ and is accompanied by vitamin deficiency. ^12^ These systemic reactions may promote coronary arteriosclerosis, leading to the occurrence and development of CHD.

However, the relationship between *H. pylori* infection and CHD is controversial. Some studies have found that *H. pylori* infection is one of the main causes of CHD, and can lead to coronary artery endothelial damage by promoting the secretion of inflammatory factors, blood coagulation, and other mechanisms, which leads to atherosclerosis and CHD, and increases the incidence rate of restenosis in patients after percutaneous transluminal coronary angioplasty (PTCA). ^13-15^ The probability of myocardial infarction in patients with *H. pylori* infection is twice that of uninfected individuals.^16^ However, some studies have found that *H. pylori* infection is not significantly related to the occurrence and degree of CHD. ^17, 18^ It is worth noting that at present, the evidence for a link between *H. pylori* infection and CHD is based on observational studies, and there may be some unknown confounding factors that affect judgment of the results. To address this controversial clinical issue, a study that removes confounding factors to accurately determine the causal relationship between *H. pylori* infection and CHD is urgently needed.

Mendelian randomization (MR) has emerged as a popular epidemiological statistical method that can remove confounding factors and accurately determine the causal relationship between two variables. The method uses the public genome-wide association study (GWAS) database to obtain instrument variables (IVs) that are strongly related to exposure but are not related to outcomes or confounding factors. The IVs are usually single nucleotide polymorphisms (SNPs), and the causal relationship between exposure and outcomes can be accurately inferred using the IVs. In this study, we used *H. pylori* infection as the exposure and applied the bidirectional MR method to infer the relationship between *H. pylori* infection and the diagnosis, prognosis, and possible pathogenesis of CHD. We also used CHD as the exposure to explore the reverse causal relationship between CHD and *H. pylori* infection, two step MR analyses were used to explore indirect pathogenic factors of *H. pylori* infection, with the aim of clarifying this relationship and providing clinical suggestions for the diagnosis and treatment of CHD.

## Methods

### Study Design

For the current study, we used IVs as a proxy for exposure, then conducted MR to test the association between exposure and outcome. ^19^ MR is based on three principle assumptions: (1) correlation assumption: IVs are strongly correlated with exposure; (2) exclusivity hypothesis: IVs are not associated with outcomes; (3) independence hypothesis: IVs are independent of other confounding factors ^20^ **(Figure 1)**.

**Figure 1.**
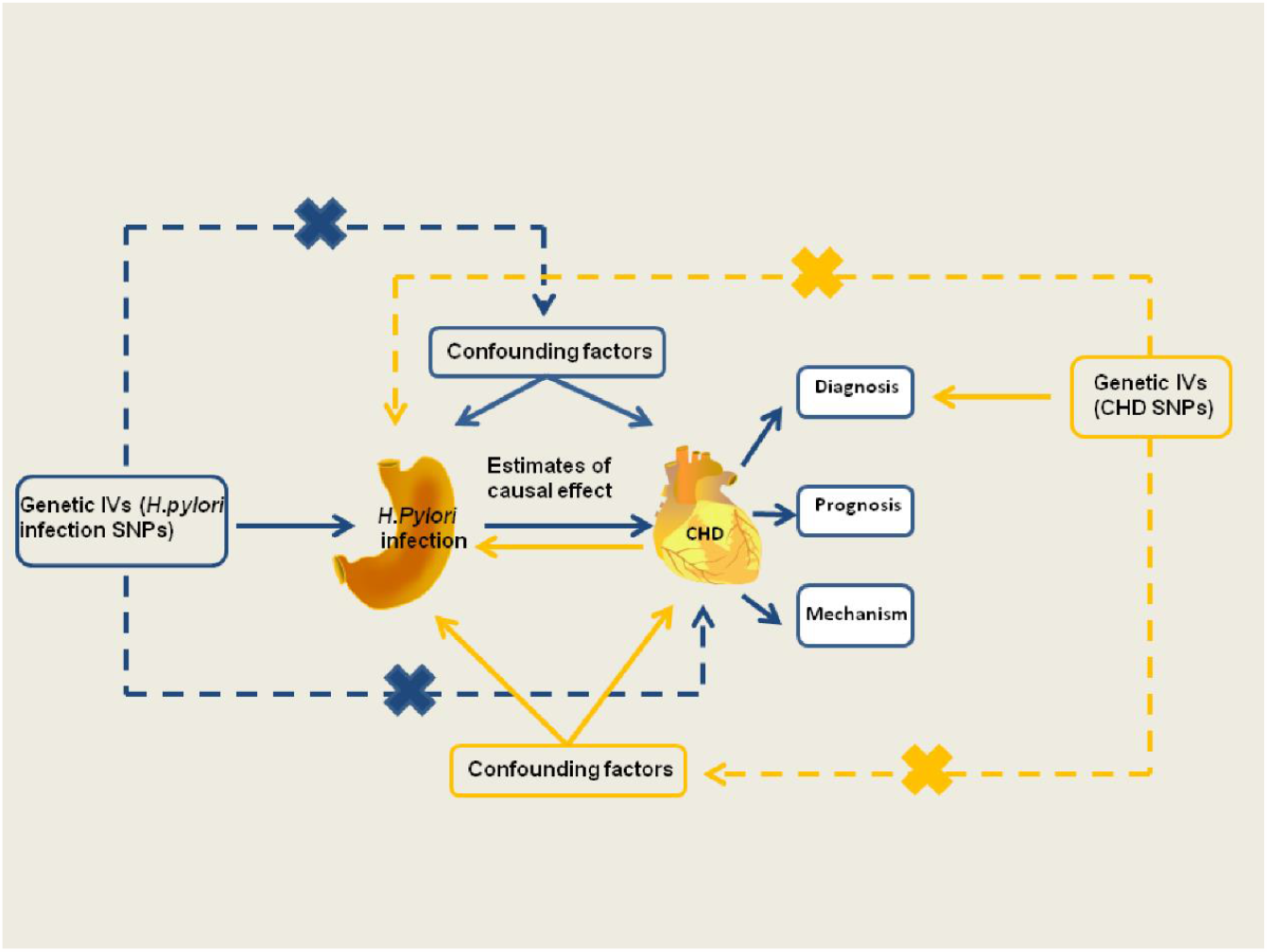
Schematic representation of the MR study on the causal relationship between *H. pylori* infection and CHD. CHD, coronary heart disease; Helicobacter pylori; IVs, instrument variants; *H. pylori*, helicobacter pylori; SNP, single-nucleotide polymorphisms.

### Data Sources Description

The genetic association of CHD was derived from the CARDIoGRAMplusC4D Consortium, which assembled 60,801 cases and 123,504 control subjects from 48 studies, and of which 77% of the participants were of European ancestry and 19% were of South and East Asian ancestry.^21^ We also collected the summary statistics for CHD, MI and angina pectoris, which were derived from the FinnGen database (https://www.fifinngen.fifi/en).^22^ *H. pylori* infection data were derived from the European Bioinformatics Institute (EBI) database (https://gwas.mrcieu.ac.uk/datasets/ieu-b-4905/) and included 1058 cases and 3625 controls. GWAS data were also collected to investigate the causal effect between *H. pylori* infection and the prognostic data for CHD, including major adverse cardiovascular events (MACE; Neale laboratory and FinnGen database), heart failure (Neale laboratory), heart arrhythmia, heart attack, stroke, target heart rate (HR) reached, and maximum HR data [MRC Integrated Epidemiology Unit (MRC-IEC), https://www.bristol.ac.uk/integrative-epidemiology]. In addition, GWAS data on the possible pathogenesis between *H. pylori* and CHD were also obtained, including fasting blood glucose data from the EBI database, body mass index (BMI) data from the MRC-IEU database, and data on lipid traits from the UK Biobank database. Vitamin data were obtained from the MRC-IEU database. Inflammation data were downloaded from the public database IEU (https://gwas.mrcieu.ac.uk/). The GWAS data are presented in detail in **Table 1** and were approved by the author or the Consortium.

**Table 1.**
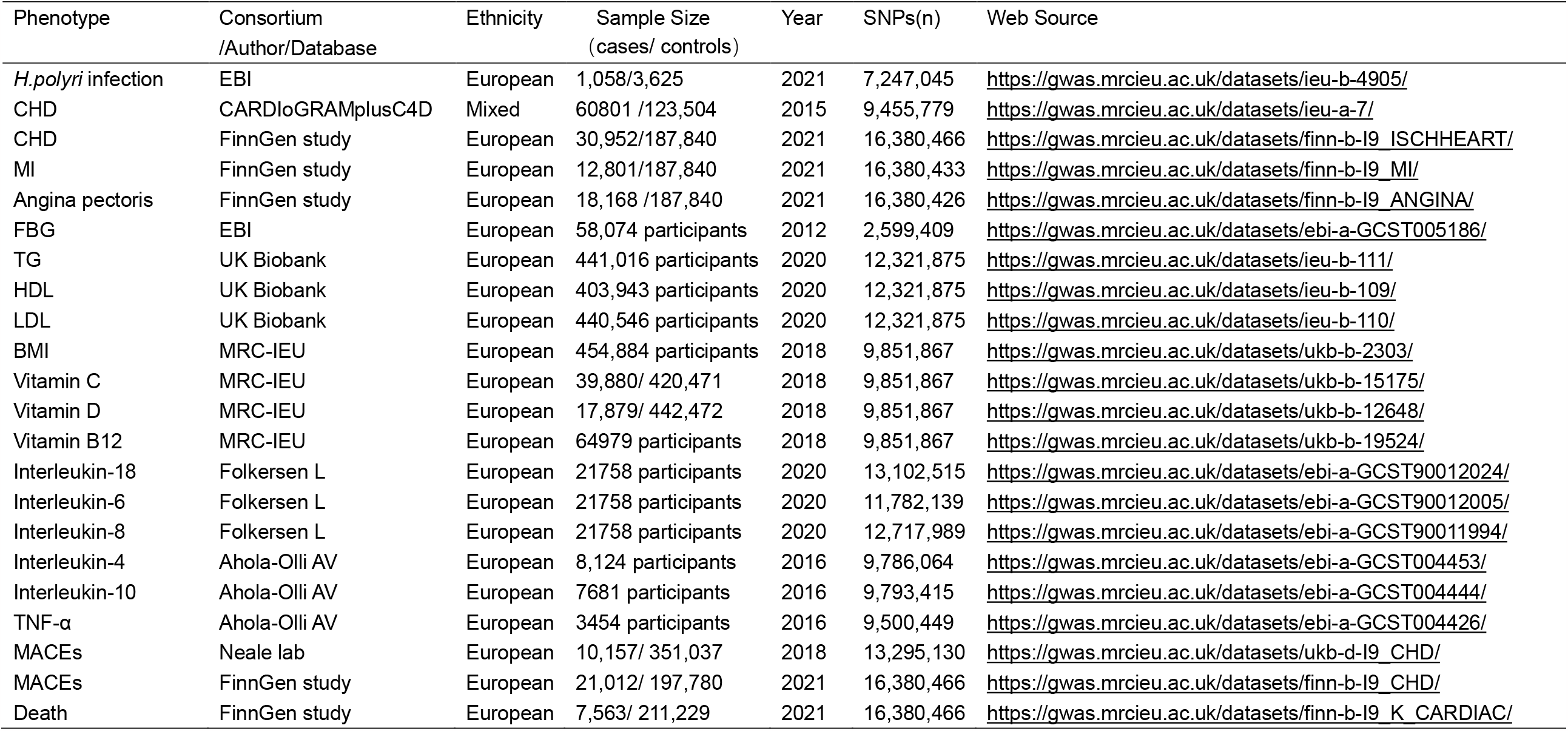
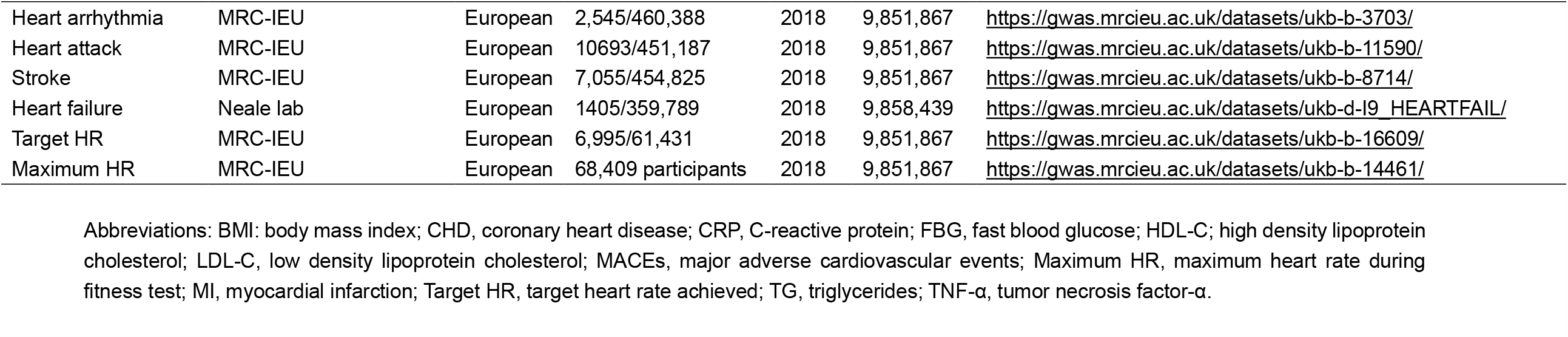
Details of the studies included in the Mendelian randomization analyses.

### Selection of Genetic Instrumental Variables for H. pylori Infection

The genetic IVs were acquired from previous literature. SNP rs368433 and SNP rs10004195 located in the Toll-like receptor 10 (*TLR10*) gene (4p14) and the Fc gamma RIIA (*FCGR2A*) gene (1q23.3), respectively, were reported to be strongly related to *H. pylori* infection as IVs^23^. Instrument strength was evaluated using the F-statistic for each allele, and if the F-statistic was greater than 10, it was considered that the potential weak instrument bias can be minimized. The F-statistic for each SNP was derived from the the following equation and was greater than 100, and could therefore be used for our analysis^24 25^ **(Table S1)**.

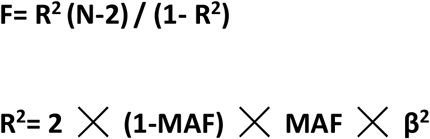

N: the sample size of the exposure dataset. R^2^: the proportion of variation explained by IVs. MAF: minor allele frequency.

### Selection of Genetic Instrumental Variables for CHD and BMI

The genetic IVs for CHD and the potential pathogenesis of *H. pylori* infection were obtained from the GWAS summary statistics. Then, the following three steps were employed to screen out the strong correlation with CHD but not that with *H. pylori* infection and confounding factors, to ensure that the effect of each allele (containing each SNP) was the same. First, SNPs strongly related to exposure were screened (*p* < 5×10^−8^). Second, independence was set to remove linkage disequilibrium (LD; r ^2^ < 0.001, window size = 10,000 kb, *p* < 5×10^−8^) and calculate the statistical strength (F-statistical > 10). Third, the exposure and outcome datasets were harmonized to ensure that the effect alleles belonged to the same allele. The SNPs screened by these strict procedures can be used as IVs for subsequent analysis **(Table S2)**. The genetic IVs for BMI were obtained by the same screening method **(Table S3)**.

### Statistical Analysis and Data Visualization

All analyses were undertaken using the R programming software (R4.1.2, https://www.rproject.org/). The primary MR analysis was conducted using the Wald ratio and the inverse variance weighted (IVW) method, and a two-sided p-value < 0.05 was considered significant, and due to the multiple comparisons, we further applied a Bonferroni corrected threshold for statistical significance (0.05/number of analyses) (**Table 2**)^26^. In the reverse MR analysis and two step MR analysis, because of the large number of IVs, we applied two complementary methods (MR-Egger and weighted-median) to increase the stability of the results. MR analyses were performed using the R-based package “TwoSampleMR” (Version 0.5.6). Forest plots were generated using the “ggplot2” R package (version 3.4.0).

**Table 2.**
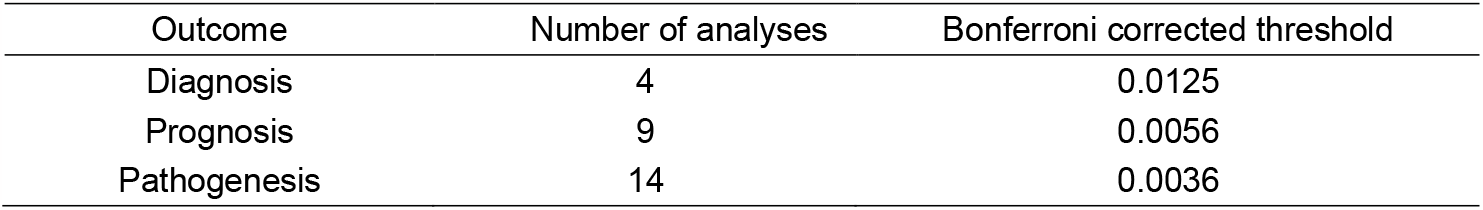
The Bonferroni corrected threshold for statistical significance in the diagnosis, prognosis and pathogenesis of CHD

| Outcome | Number of analyses | Bonferroni corrected threshold |
| --- | --- | --- |
| Diagnosis | 4 | 0.0125 |
| Prognosis | 9 | 0.0056 |
| Pathogenesis | 14 | 0.0036 |

## RESULTS

### Causal Effect of H. pylori infection on the Diagnosis of CHD

Based on previous studies, SNP rs10004195 (T > A) and SNP rs368433 (T > C) are strongly related to *H. pylori* infection, ^24^ We therefore used the two SNPs as IVs of *H. pylori* infection to predict the relationship between *H. pylori* infection and the diagnosis of CHD, MI, and angina pectoris. ^23^ Genetically predicted *H. pylori* infection showed no association with the occurrence of CHD (IEU) [odds ratio (OR), 0.991; 95% confidence interval (CI), 0.904–1.078; *p*-value = 0.842], CHD (Finn) (OR, 1.049; 95% CI, 0.980–1.118; *p*-value = 0.178), angina pectoris (OR, 1.105; 95% CI, 1.019–1.191; *p*-value = 0.023), or MI (OR, 0.993; 95% CI, 0.896–1.091; *p*-value = 0.889) under the IVW method. **(Figure 2, Table S4)**.

**Figure 2.**
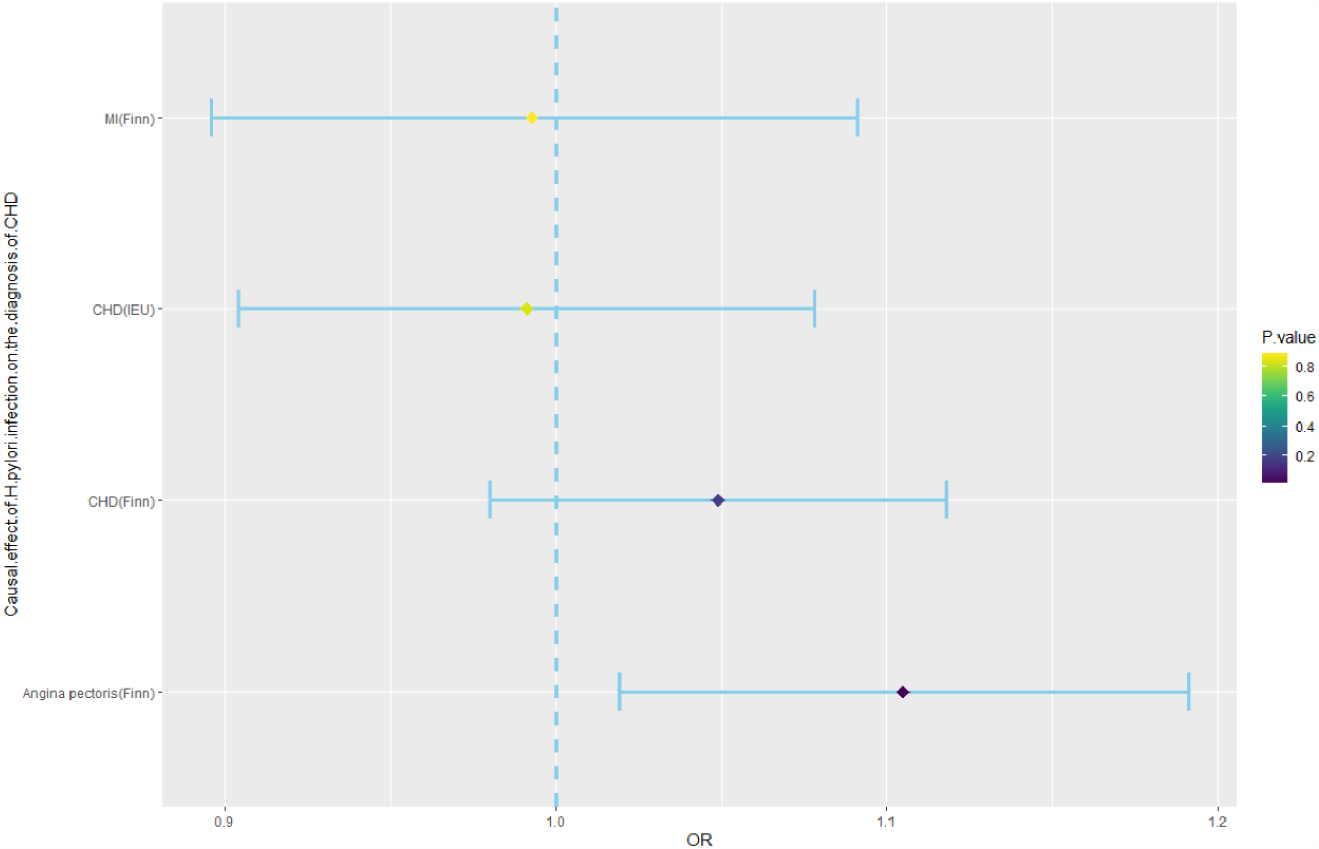
Mendelian randomization result of the effect of *H. pylori* infection on the diagnosis of CHD. CHD, Coronary heart disease; *H. pylori*, helicobacter pylori; MI, Myocardial infarction; OR, odds ratio.

### Causal Effect of H. pylori infection on the Prognosis of CHD

MR analyses were further performed to examine the causal association between *H. pylori* infection and the prognosis of CHD, including MACE, heart arrhythmia, heart attack, stroke, heart failure, target HR achieved, and maximum HR during fitness test. The analysis showed that *H. pylori* infection had no causal effect on MACE (OR, 0.999; 95% CI, 0.997–1.001; *p*-value = 0.391, IEU database; OR, 1.022; 95% CI, 0.922–1.123; *p*-value = 0.663, FinnGen database), heart arrhythmia (OR, 1.000; 95% CI, 0.999–1.001; *p*-value = 0.823), heart attack (OR, 0.998; 95% CI, 0.996–1.000; *p*-value = 0.124), stroke (OR, 0.999; 95% CI, 0.998–1.001; *p*-value = 0.525), heart failure (OR, 1.000; 95% CI, 0.999–1.001; *p*-value = 0.741), target HR achieved (OR, 0.994; 95% CI, 0.983–1.005; *p*-value = 0.252), or maximum HR (OR, 0.972; 95% CI, 0.937–1.007; *p*-value = 0.115) **(Figure 3, Table S5)**.

**Figure 3.**
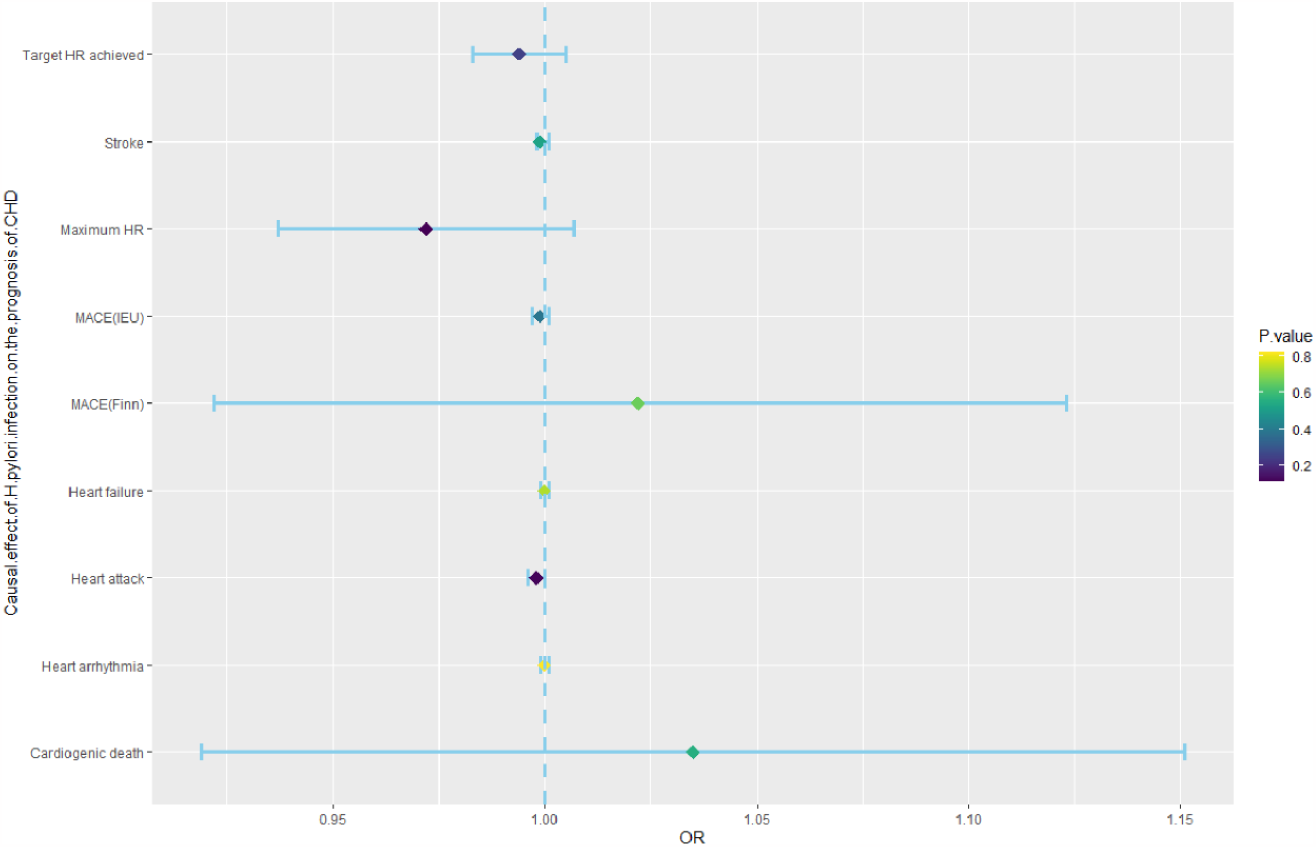
Mendelian randomization result of the effect of *H. pylori* infection on the prognosis of CHD. CHD, Coronary heart disease; *H. pylori*, helicobacter pylori; MACEs, major adverse cardiovascular events; Maximum HR, Maximum heart rate during fitness test; OR, odds ratio; Target HR achieved, reach target heart rate.

### Causal Effect of H. pylori infection on the Pathogenesis of CHD

Based on previous research, we summarized the pathogenic mechanisms of *H. pylori* infection on CHD and these included abnormal glucose and lipid metabolism, vitamin deficiency, and chronic inflammatory reaction. To explore the causal relationship between *H. pylori* infection and pathogenesis of CHD, we used *H. pylori* infection as the exposure and pathogenesis as the outcome for MR analysis. In the MR analyses of abnormal glucose and lipid metabolism, *H. pylori* infection showed no association with fasting blood glucose levels (β, 0.006; 95% CI, -0.011–0.023; *p*-value = 0.511), Triglyceride(TG) (β, 0.005; 95% CI, -0.006–0.016; *p*-value = 0.409), High-density lipoprotein cholesterol (HDL-C) (β, -0.006; 95% CI, -0.047–0.035; *p*-value = 0.788), or low-density lipoprotein cholesterol (LDL-C) (β, 0.013; 95% CI, -0.026–0.051; *p*-value = 0.515). In the MR analysis of vitamin deficiency, we also obtained negative results, including vitamin C (β, -0.002; 95% CI, -0.006–0.002; *p*-value = 0.318), vitamin D (β, -0.0003; 95% CI, -0.003–0.002; *p*-value = 0.775), and vitamin B12 (β, 0.008; 95% CI, -0.029–0.044; *p*-value = 0.685). However, there was a significant causal effect between *H. pylori* infection and BMI (β, 0.022; 95% CI, 0.008–0.036; *p*-value = 0.001), and there is a causal relationship between BMI and CHD. In the analyses of an inflammatory reaction, no positive results were obtained for interleukin families or tumor necrosis factor **(Figure 4, Table S6, and Table S7)**.

**Figure 4.**
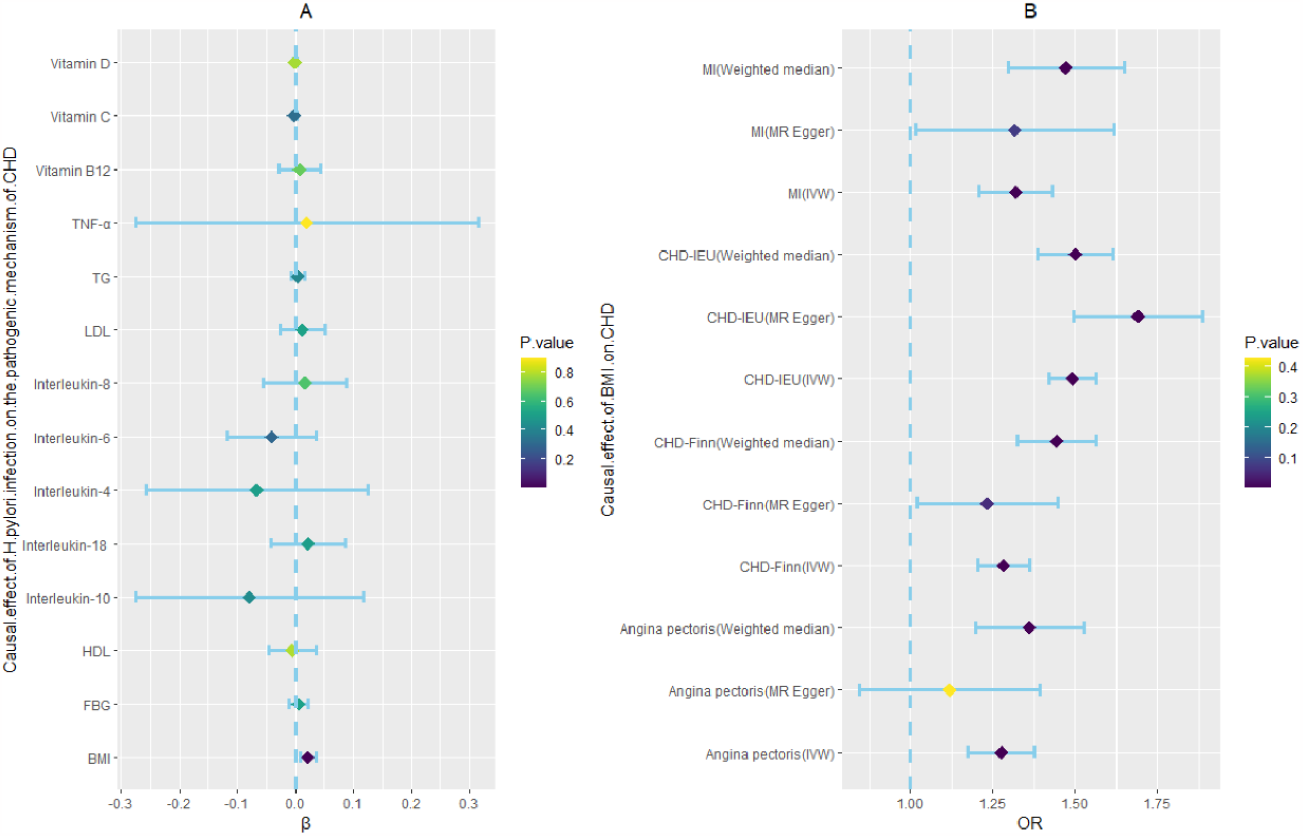
Two step Mendelian randomization result of the effect of *H. pylori* infection on CHD (the pathogenic mechanism of CHD). BMI: body mass index; CRP, C-reactive protein; FBG, fast blood glucose; *H. pylori*, helicobacter pylori; HDL-C, high density lipoprotein cholesterol; LDL-C, low density lipoprotein cholesterol; OR, odds ratio. TG, triglycerides; TNF-α, tumor necrosis factor-α. Dark blue dots represent statistical differences in *P* values.

### Reverse Causal Effect of CHD on H. pylori Infection

The IVs of CHD, MI, and angina pectoris were identified from the public GWAS summary data. Three MR analysis methods, namely, inverse variance weighted (IVW), weighted median, and MR-Egger, were used for this analysis. All three methods showed no significant causal relationship on *H. pylori* infection **(Figure 5, Table S8)**.

**Figure 5.**
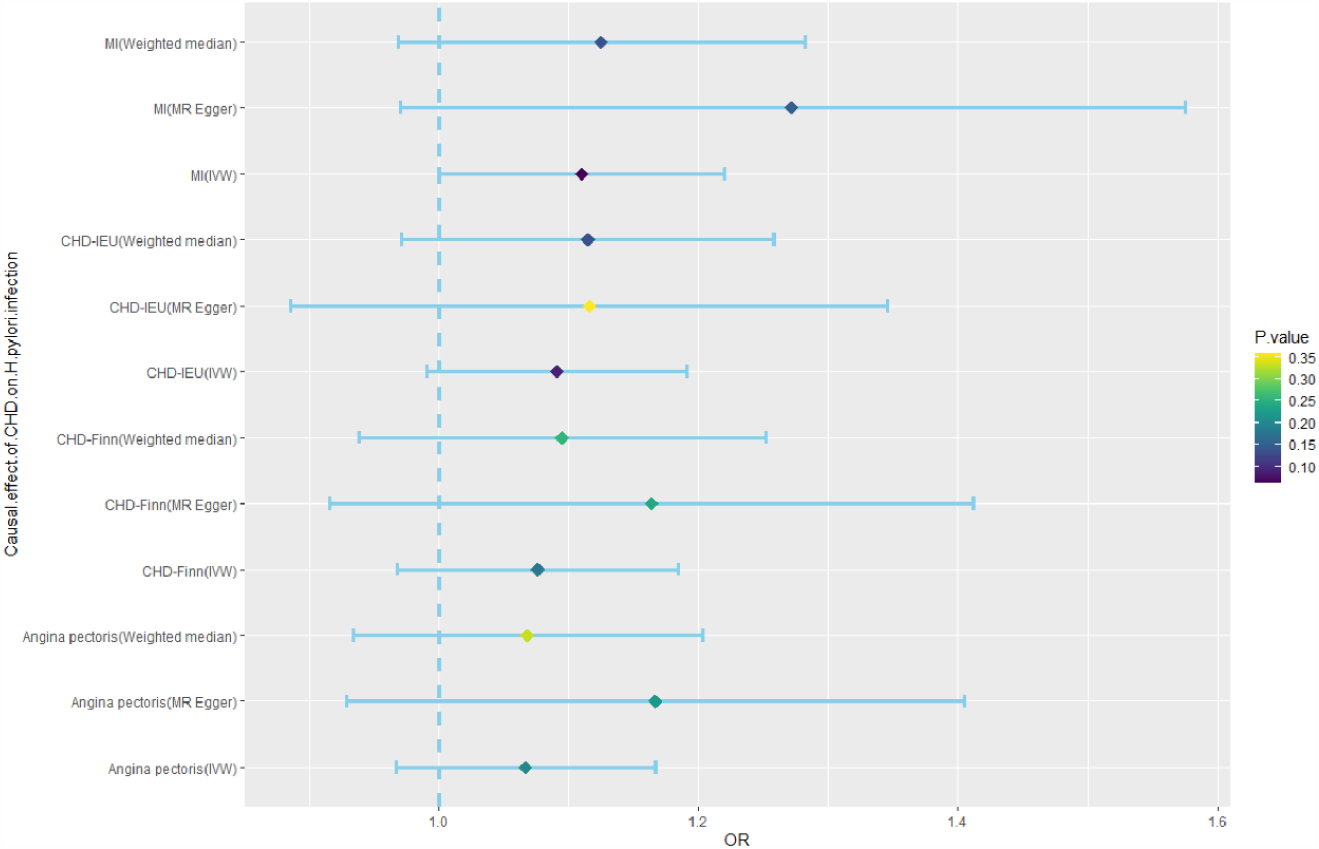
Mendelian randomization result of the effect of CHD on *H. pylori* infection. CHD, coronary heart disease; *H. pylori*, helicobacter pylori; MI, myocardial infarction; OR, odds ratio.

## Discussion

In this study, we used large-scale public GWAS data to analyze the causal relationship between *H. pylori* infection and the risk of CHD using the MR method. The causal effect of *H. pylori* infection on CHD is mediated by BMI.

The association between *H. pylori* infection and CHD is currently controversial. Several studies have reported that *H. pylori* infection is related to the occurrence and prognosis of CHD. ^15, 27, 28^ A prospective study found that *H. pylori*-infected patients had an increased occurrence of CHD ^29^ and adverse events.^27^ In other studies, MI patients infected with *H. pylori* had a higher mortality rate ^30^ and the probability of restenosis after PTCA was higher.^13^ It has also been shown that MI has a reverse causal effect on *H. pylori*. Young people with MI have double the probability of *H. pylori* infection as healthy individuals. ^31^ However, some studies have been unable to detect a correlation between the occurrence and development of CHD and *H. pylori* infection, especially among older individuals. ^32^ A prospective study on a small sample ^18^ and meta-analyses of five large samples ^18, 33^ have all provided evidence that *H. pylori* infection is not significantly related to the severity and prognosis of CHD. A prospective study involving 180 patients found that *H. pylori* infection is not significantly related to restenosis after PTCA. ^34^ The pathogenic link between *H. pylori* infection and CHD remains controversial. First, in terms of metabolism, the influence of *H. pylori* infection on glucose and lipid metabolism and BMI is controversial. With regard to lipids, a study found that *H. pylori* infection can reduce the level of HDL and increase the levels of LDL and TG.^35^ However, other studies present the opposite findings. Meta-analyses and prospective studies of large samples have shown that eradication of *H. pylori* infection has no significant effect on the levels of HDL, TG, and LDL.^6, 36^ In terms of glucose metabolism, evidence suggests that *H. pylori* infection may participate in the onset of diabetes and impaired glucose control in diabetes patients.^37, 38^ Patients with *H. pylori* infection from different populations also reported higher levels of insulin resistance.^39^ One study found that compared with the control group, the improvement in glucose homeostasis in diabetes patients after successful eradication of *H. pylori* infection was not statistically significant.^40^ With regard to weight, eradication of *H. pylori* infection may increase the weight of children ^41^ and increase ^42^ or decrease ^43^ the weight of adults, and obese individuals show a higher prevalence of *H. pylori* infection^44^. Second, *H. pylori* infection causes changes in the gastrointestinal microenvironment, which may impair the absorption of nutrients, leading to a lack of micronutrients. Poor vitamin absorption has been shown to be related to *H. pylori* infection^45^. Vitamin C and D levels are closely related to CHD, but the relationship between vitamins and *H. pylori* remains to be confirmed. Third, *H. pylori* can cause an inflammatory reaction. Chronic inflammation caused by *H. pylori* may have dual effects. On the one hand, low-grade inflammation is a common feature of obesity, diabetes, insulin resistance, and dyslipidemia and *H. pylori* may cause a chronic inflammatory reaction through abnormal metabolism.^46^ On the other hand, *H. pylori* causes damage to the gastrointestinal tract,^47^ stimulating the increase in IL levels ^10, 11^. *H. pylori* infection has been associated with elevated levels of tumor necrosis factor and IL-6 in patients with CHD.^48^ Conflicting data has also been reported regarding inflammation. ^49^ The factors involved in the pathogenesis of *H. pylori* infection, which include glucose and lipid metabolism, vitamin deficiency, and chronic inflammatory reactions, are all causes of CHD.

The controversy between *H. pylori* infection and CHD could be attributed to multiple reasons, such as differences in the race and age of the selected sample population, the small sample size, the low incidence of MACE, the detection method for *H. pylori* infection, and the different follow-up times. These confounding factors may lead to the poor statistical efficiency of the data and may affect the reliability of the experimental results.

This study found that *H. pylori* infection has no direct causal effect on the diagnosis and prognosis of CHD. In the analysis of pathogenesis, *H. pylori* infection has a causal effect on BMI, and BMI has a causal effect on CHD. Therefore, the causal effect of *H. pylori* infection on CHD is mediated by BMI. However, *H. pylori* infection has no causal effect on inflammatory factors, vitamins, glucose and lipid metabolism and there is no reverse causal effect of CHD on *H. pylori* Infection.

This study used the MR method to reveal a bidirectional causal relationship between *H. pylori* infection and CHD for the first time, and could increase the recognition of pathogenic factors of CHD from the perspective of system biology. The advantage of the MR study is that the sample size is large, and it is a natural randomized controlled trial, which eliminates confounding factors. However, there are still limitations of this study. The GWAS of *H. pylori* infection is based on serological samples, which may not be truly representative of *H. pylori* infection. Furthermore, the samples were of european origin and therefore may not be representative of populations worldwide. Finally, the screening of IVs in this study was strict, which may have led to negative results.

## Conclusions

Our findings confirm that the causal effect of *H. pylori* infection on CHD is mediated by BMI. Therefore, eradication or prevention of *H. pylori* infection may have a clinical benefit for patients with CHD indirectly. However, it is not recommended that patients with CHD be screened for *H. pylori* infection.

## Data Availability

The datasets presented in this study can be downloaded from the public website (Table 1)

## Nonstandard Abbreviations and Acronyms

CHD: coronary heart disease
GWAS: genome-wide association studies
H. pylori: helicobacter pylori
IVs: instrument variables
IVW: inverse variance weighted
MACE: major adverse cardiovascular events
MR: mendelian randomization
SNPs: single nucleotide polymorphisms

## Acknowledgement

We thank the personnel at our centers participating in the research for their commitment and professionalism to this study.

## Author Contributions

Bing Li, Yang Zheng, and Yaoting Zhang analyzed the study data, Bing Li wrote the manuscript. He Cai and Yang Zheng provided ideas and supervised the research and edited the manuscript. All authors approved the submitted version.

## Data Availability

The datasets presented in this study can be downloaded from the public website (Table 1).

## Supplemental Material

Tables S1-S8

